# Comparing the efficacy and safety of direct oral anticoagulants versus Vitamin K antagonists in patients with antiphospholipid syndrome: a systematic review and meta-analysis

**DOI:** 10.1101/2022.04.11.22273703

**Authors:** Xiaoling Wu, Shaobo Cao, Bo Yu, Tao He

## Abstract

**Background:** Thromboprophylaxis is the cornerstone strategy for thrombotic antiphospholipid syndrome (APS). Data comparing direct oral anticoagulants (DOACs) to Vitamin K antagonists (VKAs) in the secondary prevention of thrombosis in APS patients remain contentious.

**Objectives:** We aim to review and analyze literature on the efficacy and safety of DOACs compared to VKAs in treating patients with APS. A literature search was performed from inception to March 1, 2022. Subgroups were analyzed based on the risk stratification of APS profiles and different DOAC types.

**Results:** A total of 9 studies with 1131 patients were included in the meta-analysis. High-risk APS patients (triple positive APS) who used DOACs displayed an increased risk of recurrent thrombosis (RR=3.65, 95% CI:1.49-8.93; I^2^=29%, P=0.005) compared to those taking VKAs. Similar risk of recurrent thrombosis or major bleeding was noted in low-risk APS patients (single or double antibody-positive) upon administering DOACs or VKAs. The utilization of Rivaroxaban was associated with a high risk of recurrent thromboses (RR=2.63; 95% CI, 1.56-4.42; I^2^ =0, P=0.0003), particularly recurrent arterial thromboses (RR=4.52; 95% CI, 1.99-10.29; I^2^ =0, P=0.18) in overall APS patients. Comparisons of the rate of recurrent thrombosis events and major bleeding events when using dabigatran or apixaban versus VKAs yielded no statistical differences.

**Conclusions:** In the absence of contraindications, this meta-analysis suggests that VKAs remain the first-choice treatment for high-risk APS patients, with DOACs a more appropriate option for low-risk APS patients. Different DOACs may exhibit different levels of efficacy and safety for thromboprophylaxis in APS patients and require further exploration.

## Introduction

Antiphospholipid syndrome (APS) is an acquired autoimmune disorder manifested by the persistent presence of antiphospholipid antibodies (aPL) (including lupus anticoagulant (LA), anticardiolipin antibodies (aCL) and anti-β2 glycoprotein I antibodies (aβ2-GPI)), which can result in recurrent thrombophilia and obstetrical morbidity. [1] It is clinically heterogeneous, with the risk of occurrence and recurrence of thrombosis dependent on the profile of aPL, concomitant diseases, and anticoagulation strategies. [2] Thromboprophylaxis is the absolute cornerstone strategy for thrombotic APS; however, controversies exist at the level of anticoagulant drug selection.

Though the use of Vitamin K antagonists (VKAs), notably warfarin (target international normalized ratio [INR], 2 to 3), requires that patients are highly compliant to a healthy lifestyle and that INR is monitored frequently, they remain the recommended standard therapy for long-term thromboprophylaxis in patients with APS after the first thrombotic episode. [3] Compared to VKAs, direct oral anticoagulants (DOACs, such as rivaroxaban, apixaban, edoxaban and dabigatran) have appealing benefits, including no need to monitor anticoagulants’ effect, fewer drug-food interactions, fixed-dose prescribing, fewer cases of significant bleeding, and many more. [3] To date, data comparing DOACs to VKAs for the secondary prevention of thrombosis in APS patients remain limited and debatable. Of the published cohort studies and randomized clinical trials (RCTs) related to thromboprophylaxis in APS patients, some found that DOACs were not substandard to VKAs, [4-7] and the others produced contrasting results. [8-13]

Four reviews and meta-analyses evaluating the efficacy and safety of DOACs versus VKAs in APS patients have been published. Two of these investigations have limited value presently because cohort studies and RCTs were rarer at the time of their completion than they are now; [14,15] cohort evaluations and RCTs remain incredibly scarce. Two related meta-analyses were published recently: one included 4 RCTs, [16] and the other enrolled 3 RCTs and 4 observational studies. [17] Dufrost et al.’s assessment of 4 RCTs concluded that the DOACs used in APS patients were not less effective than VKAs in preventing recurrent venous thromboembolism; however, they significantly increased the risk of recurrent arterial thrombosis. [16] On the other hand, Koval demonstrated that DOACs, particularly rivaroxaban, provided poorer efficiency than VKAs when given to patients with APS because they tripled the thromboembolic risk. [17]

Evidence from case series, RCTs, and a meta-analysis has indicated that high-risk APS patients with triple positivity or a history of arterial thrombosis are associated with a higher risk of thrombosis. [8-10,12,18] Several international guidelines do not recommend DOACs in place of warfarin (INR goal 2–3) as preferred thromboprophylaxis treatment for high-risk APS patients. [1,3,19] However, whether and which DOACs can be used in low-risk APS patients (presence of one or two antiphospholipid antibody profiles) warrants further exploration given their advantages. Both meta-analyses mentioned above did not perform subgroup evaluations based on different risk stratifications of APS or detailed DOAC drug types. [16,17]

Three retrospective cohort studies on the subject were also published recently. [6,7,12] One was performed upon the completion of the Antiphospholipid Syndrome (TRAPS) Trial, with most patients involved in the TRAPS trial switched to warfarin, except for six patients who remained on DOACs. [12] Interestingly, the subjects enrolled in the two latest cohort studies were mostly low-risk APS patients; still, both investigations yielded contradictory results. [6,7] In light of that outcome and existing controversies, we conducted this systematic review and meta-analysis to compare the efficacy and safety of DOACs versus VKAs for thromboprophylaxis in different risk-stratified APS patients. We also sought to establish whether different DOACs display varying levels of efficacy and safety for thromboprophylaxis in APS patients.

## Methods

### Search strategy and study selection

We systematically searched PubMed, the Cochrane Central Register of Controlled Trials and EMBASE using the following keywords and their Mesh terms: “antiphospholipid syndrome” and “direct oral anticoagulants”, “novel oral anticoagulant” or “apixaban”, “dabigatran”, “edoxaban”, and “rivaroxaban” until March 1, 2022. Search language was restricted to English. References in identified reports and review articles were also mined to identify potentially relevant studies. This research was performed per the Preferred Reporting Items for Systematic Reviews and Meta-Analyses (PRISMA) guidelines.

### Inclusion and exclusion criteria

Studies comparing reported clinical outcomes in patients with APS using DOACs or VKAs were included. The following types of articles were excluded: review articles or commentaries, case reports, and articles not in the English language. Investigations with insufficient data for estimating the efficacy and safety of DOACs were also excluded. Multiple articles published by the same institution and with overlapping sbjects were scrutinized, and that with the most significant number of cases or the most extended follow-up was selected; the others were excluded.

### Data Extraction and quality of study assessment

Two reviewers independently extracted data from each article into a pre-specified data collection form. Materials were mined from the main text and tables of the published reports and online supplementary materials (if available). If available, the following items were retrieved from each publication: the first author’s name, publication year, design and setting, number of subjects, anticoagulation treatment regimen, patient characteristics, follow-up period, and clinical outcomes. The outcome of interest was thromboembolic events, including venous and arterial thromboembolism, major bleeding and all-cause mortality. Major bleeding was diagnosed in each study using guidelines from the International Society on Thrombosis and Haemostasis. [20]

The risk of biased assessments of RCTs was conducted in line with the criteria in the Cochrane Handbook for Systematic Reviews of Interventions. [21] This methodology explores the adequacy of sequestration, allocation sequence concealment, blinding of participants and study personnel, blinding for outcome assessment, incomplete outcome or selective outcome reporting, and other potential biases. The quality of the included cohort studies was modified evaluated using the Newcastle-Ottawa scale (NOS).[22] The two reviewers independently analyzed the selection, comparability, and exposure of each publication and allocated a score of between 0-9 to each included cohort. Studies with scores ≥ 7 were considered suitable for analysis. Any disagreement between the authors was resolved via dialog with a senior reviewer.

### Statistics

Data were managed and analyzed using Review Manager Software (version 5.4; the Cochrane Collaboration) and STATA software (version 12.0; STATA Corporation, College Station). The Mantel-Haenszel method for dichotomous data was used to calculate aggregated risk ratios (RRs) and corresponding 95% confidence intervals (CIs). Results were considered statistically significant at P-values <0.05. Unexplained statistical heterogeneity between studies was assessed with the aid of the I^2^ statistic. A fixed-effects model was used at I^2^ ≤ 50% and Cochran Q statistic P > 0.1, and a random-effects model was applied at I^2^ > 50% and Q statistic P ≤ 0.1. Subgroup analyses were conducted based on the study designs (RCT and cohort study), DOAC types (rivaroxaban, apixaban, dabigatran), and different risk stratifications of APS (triple antibody-positive, single or double antibody-positive) to avoid method heterogeneity. APS patients with triple-antibody positivity or a history of arterial thrombosis are generally defined as high-risk, while those with single- or double-antibody positivity are low-risk. Sensitivity was analyzed by excluding one study after the other and re-analyzing data. Publication bias was evaluated using Begg’s test and Egger’s test, with significant publication bias considered existent at P-values <0.05. [23,24]

## Results

### Search Results and the Characteristics of Included Trials

Figure 1 shows the PRISMA flow chart summarizing the search strategy. A total of 9 articles, including 4 RCTs and 5 cohort studies, with 1131 patients from the literature search were included in this meta-analysis. The RAPS[4], TRAPS[9] and EUDRA[10] were open-label and non-inferiority trials. Goldhaber et al.’s inquiry examined APS patients with previous VTE in RE-COVER®, RE-COVER II™ and RE-MEDY™ double-blind, randomized controlled trials comparing dabigatran with warfarin. [5] Four of the five cohort studies we selected were retrospective, and one was prospective in design. Peng’s retrospective cohort research, which switched the treatment regimen of most subjects in the DOACs group from the TRAPS trial to warfarin, was excluded because of its association to that trial that ended just before its run. [12] The characteristics of the RCTs and cohort studies are described in full in Table 1.

**Table 1.** Characteristics of included studies.

| First author | Year | Design | Criteria | Exclusion | Country | Relevant patients |  | Age, mean±SD, or median |  | Triple |  | Intervention |  | Follow-up |  |
| --- | --- | --- | --- | --- | --- | --- | --- | --- | --- | --- | --- | --- | --- | --- | --- |
|  |  |  |  |  |  | DOAC | VKA | DOAC | VKA | DOAC | VKA | DOAC(n) | VKA | DOAC | VKA |
| Benjamin Franke | 2020 | Retrospective cohort | High risk and low risk | NR | Germany | 119 | 81 | 55 (21 - 81) | 51 (21-76) | 7% | 4.2% | Rivaroxaban 20mg/d (92), apixaban 5mg twice daily (42), dabigatran 150mg twice daily (3) and edoxaban 60mg/d (4) | INR 2.0-3.0 | 16 months | 32 months |
| Briana Williams | 2021 | Retrospective cohort | Low risk | Triple positive, history of arterial thrombosis | USA | 39 | 57 | 56 (39-66) | 54 (36-69) | 0 | 0 | Rivaroxaban (35), dabigatran (3), apixaban (1) | INR 2.0-3.0 | NR | NR |
| Goldhaber | 2016 | RCT | High risk and low risk | NR | International | 71 | 80 | 47.8 (14.9) | 47.4 (16.8) | NR | NR | Dabigatran 150mg twice daily (71) | INR 2.0-3.0 | NR | NR |
| Hannah Cohen | 2016 | RCT | High risk and low risk | history of arterial thrombosis, recurrent venous thromboembolism when taking VKA, pregnancy, severe renal impairment, thrombocytopenia | UK | 57 | 59 | 47 (17) | 50 (14) | 12% | 20% | Rivaroxaban 20mg/d (or 15mg/d for patients with a creatinine clearance of 30 to 49 mL/min/1.73 m<sup>2</sup>) (57) | INR 2.0-3.0 | 210 days | 210 days |
| Ida Martinelli | 2018 | Retrospective cohort | High risk and low risk | NR | Italy | 13 | 15 | 46.2 (16.4) | 43.1 (15.8) | 46.2% | 46.7% | Rivaroxaban (15mg twice daily for 21 days followed by 20 mg/day) (13) | INR 2.0-3.0 | 43 months | 48 months |
| Malce K | 2020 | Prospective cohort | High risk and low risk | Stroke within the previous 3 months, pregnancy, severe renal impairment, thrombocytopenia | Poland | 82 | 94 | 44±11 | 45±13 | 40% | 21% | Rivaroxaban 20mg/d (36), apixaban 5mg twice daily (42), dabigatran 150mg twice daily (4) | INR 2.0-3.0 | 45 months | 62 months |
| Ordi-Ros | 2019 | RCT | High risk and low risk | Had intracranial hemorrhage, stroke, or gastrointestinal bleeding within the previous 3 months, significant bleeding diathesis, pregnancy, severe renal impairment, thrombocytopenia | Spain | 95 | 95 | 47 (40 - 55) | 51 (38-63) | 61% | 60% | Rivaroxaban 20mg/d (or 15mg/d for patients with a creatinine clearance of 30 to 49 mL/min/1.73 M<sup>2</sup>) (95) | INR 2-3 (or 3.1-4.0 if history of recurrent thrombosis) | 34.1 months | 33.1 months |
| Pengo | 2018 | RCT | High risk | Significant bleeding diathesis, current pregnancy or breast feeding, severe renal impairment, concomitant treatment with other anticoagulants | Italy | 59 | 61 | 46.5 ±10.2 | 46.1 ±13.2 | 100% | 100% | Rivaroxaban 20mg/d (or 15mg/d for patients with a creatinine clearance of 30 to 49 mL/min/1.73 M<sup>2</sup>) (59) | INR 2.0-3.0 | 611 days | 611 days |
| Sato T | 2019 | Retrospective cohort | High risk and low risk | NR | Japan | 18 | 39 | 47.7 ±17.1 | 42.6 ±13.4 | 33.3% | 38.9% | Rivaroxaban (5), edoxaban (12), apixaban (1) | INR 2.0-3.0 | 60 months | 60 months |

NR: not reported, VKA: Vitamin K antagonist, DOAC, direct oral anticoagulant

**Fig. 1.**
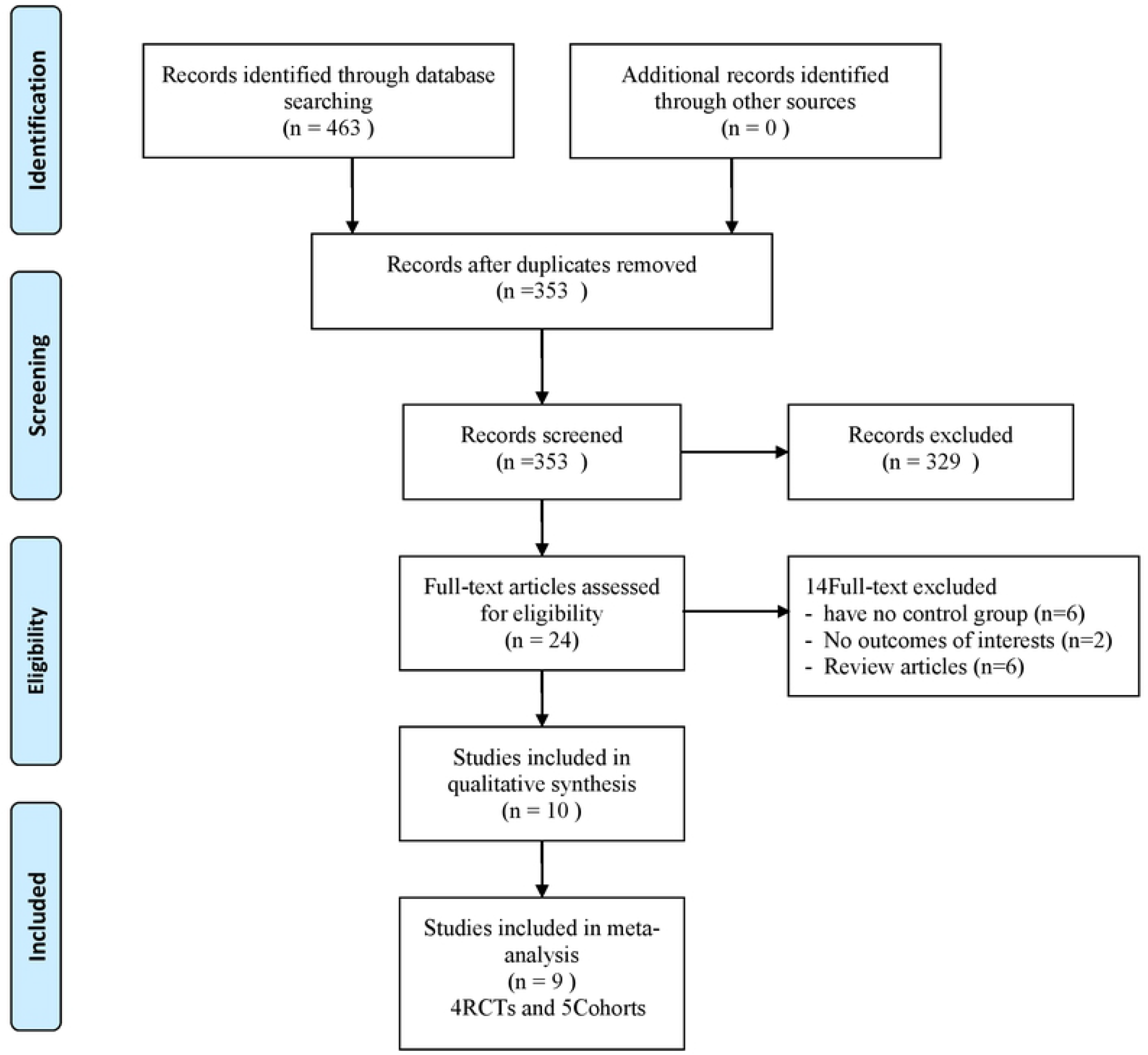
Flow chart on the selection of eligible studies

### Quality of study assessment

The risk of bias assessment for the included RCTs is presented in Fig 2. The performance biases for three trials with open-label study designs[4,9,10] were ambiguous, as were the selective reporting bias for Goldhaber et al.’s investigation[5] because it was a post hoc analysis of three previous published RCTs and the follow-up duration time bias in the RAPS trial due to the relatively short follow-up time. The selection, detection, and attrition biases were deemed low risk for all trials. In summary, all the RCTs were considered high-quality.

**Fig. 2.**
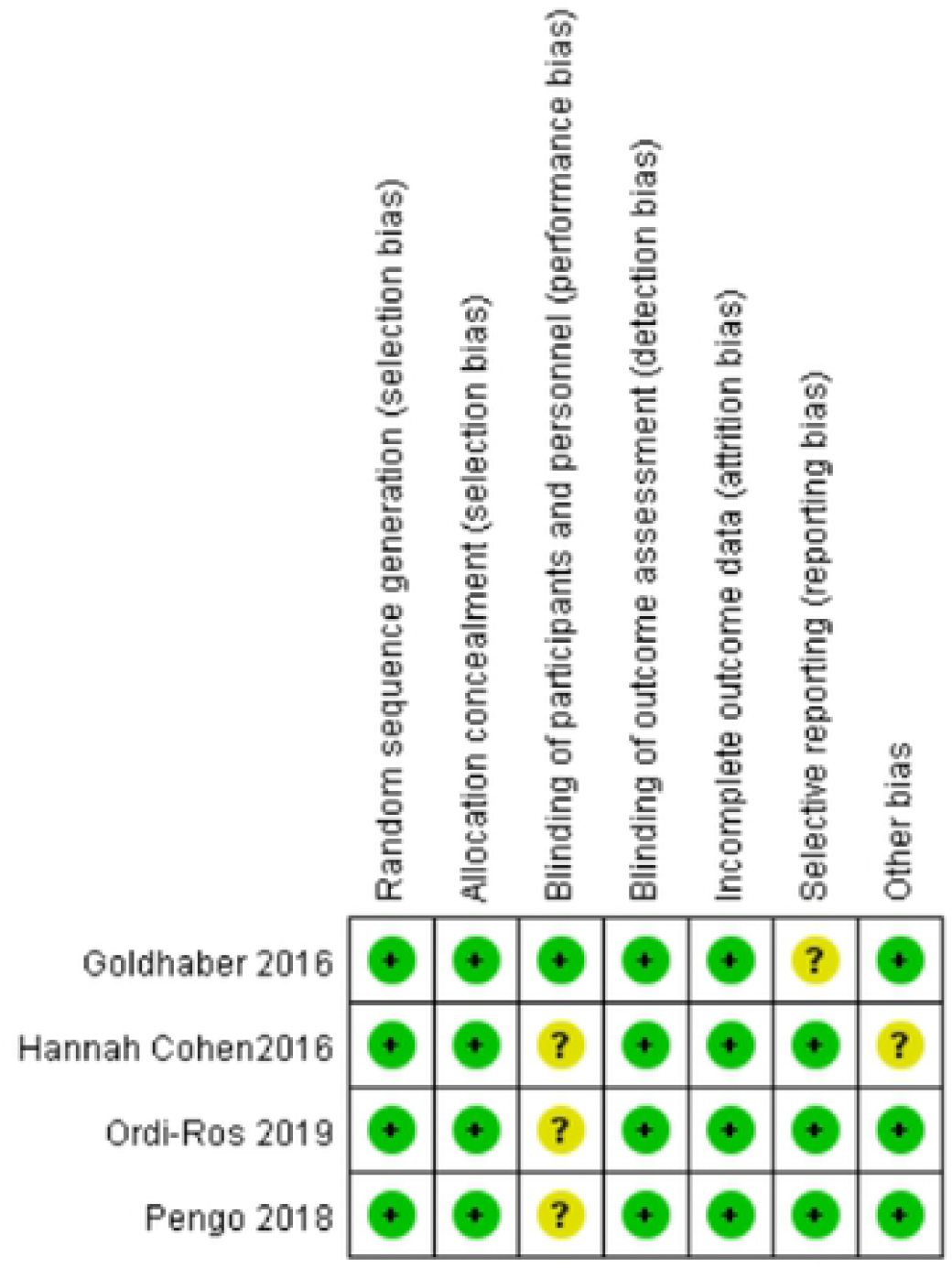
Risk of bias assessment for RCTs

The risk of bias assessment for the cohort studies was assessed with NOS (Supplementary Table 1). All selected studies scored ≥7 and were deemed fit for inclusion in the meta-analysis.

### Clinical outcomes

#### Comparing the efficacy and safety of DOACs versus VKAs in overall APS patients (Fig 3)

First, we determined the pooled RR of DOACs relative to VKAs for thromboprophylaxis in overall APS patients. There was no significantly increased risk of recurrent thrombosis (RR=1.53, 95% CI: 0.92-2.55; I^2^=24%, P=0.10) (Fig 3A) and recurrent venous thromboembolism (RR=1.22, 95% CI: 0.68-2.17; I^2^=0%, P=0.51) in the DOAC group (Fig 3B) compared to the VKA group in both RCTs and cohort studies, as determined by subgroup analyses (Fig 3A&B). Disappointingly, DOACs showed a markedly considerable aptitude for risk of recurrent arterial thrombosis (RR=2.27, 95% CI: 1.28-4.00; I^2^=29%, P=0.005) in APS patients (Fig 3C). The RCT subgroup displayed a substantially more inflated risk of recurrent arterial thrombosis (RR=5.33, 95% CI: 1.74-16.32; I^2^=0%, P=0.003), while the subgroup of cohort studies exhibited no noteworthy differences (RR=1.36, 95% CI: 0.68-2.71; I^2^=9%, P=0.38) (Fig 3C).

**Fig. 3.**
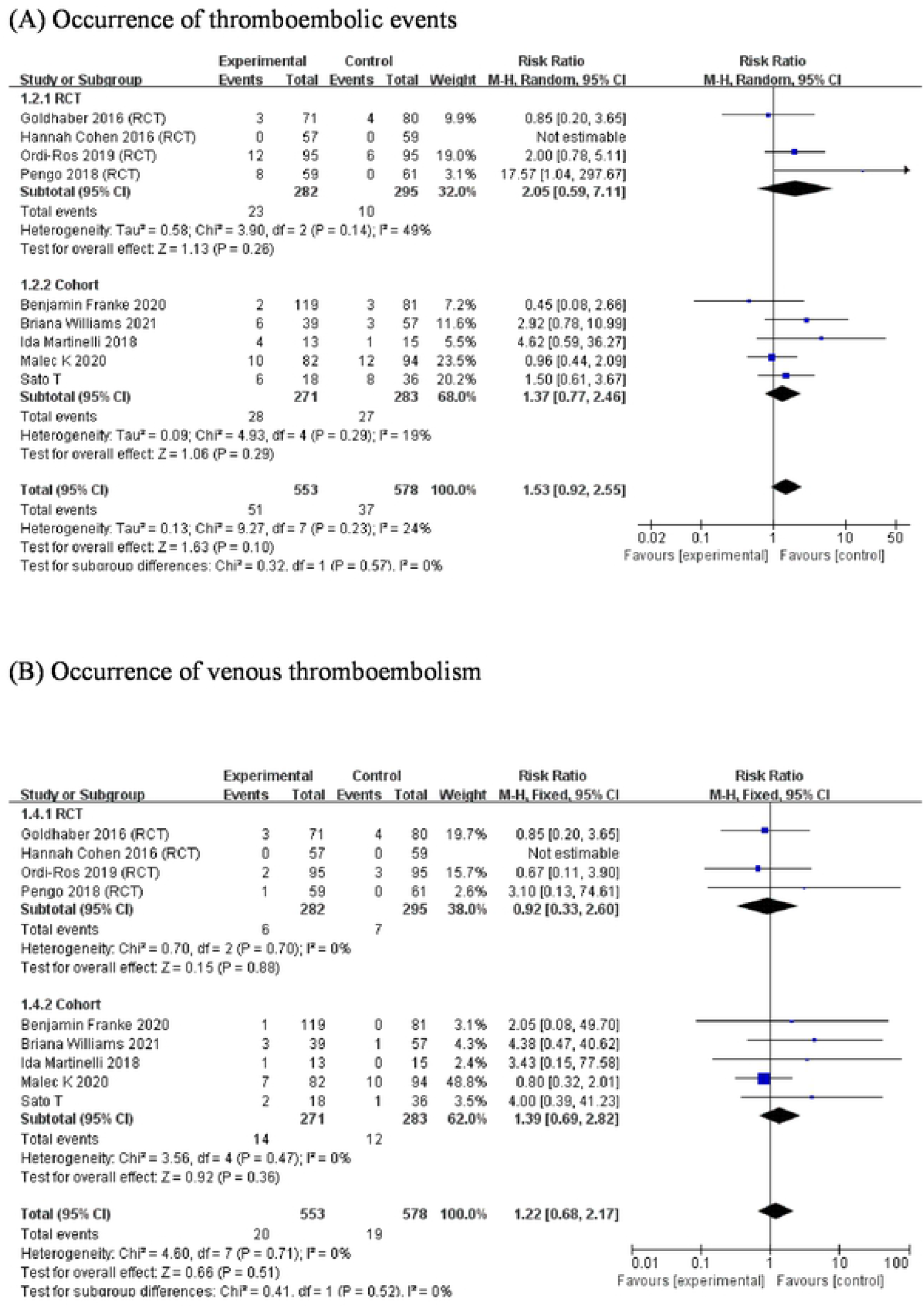
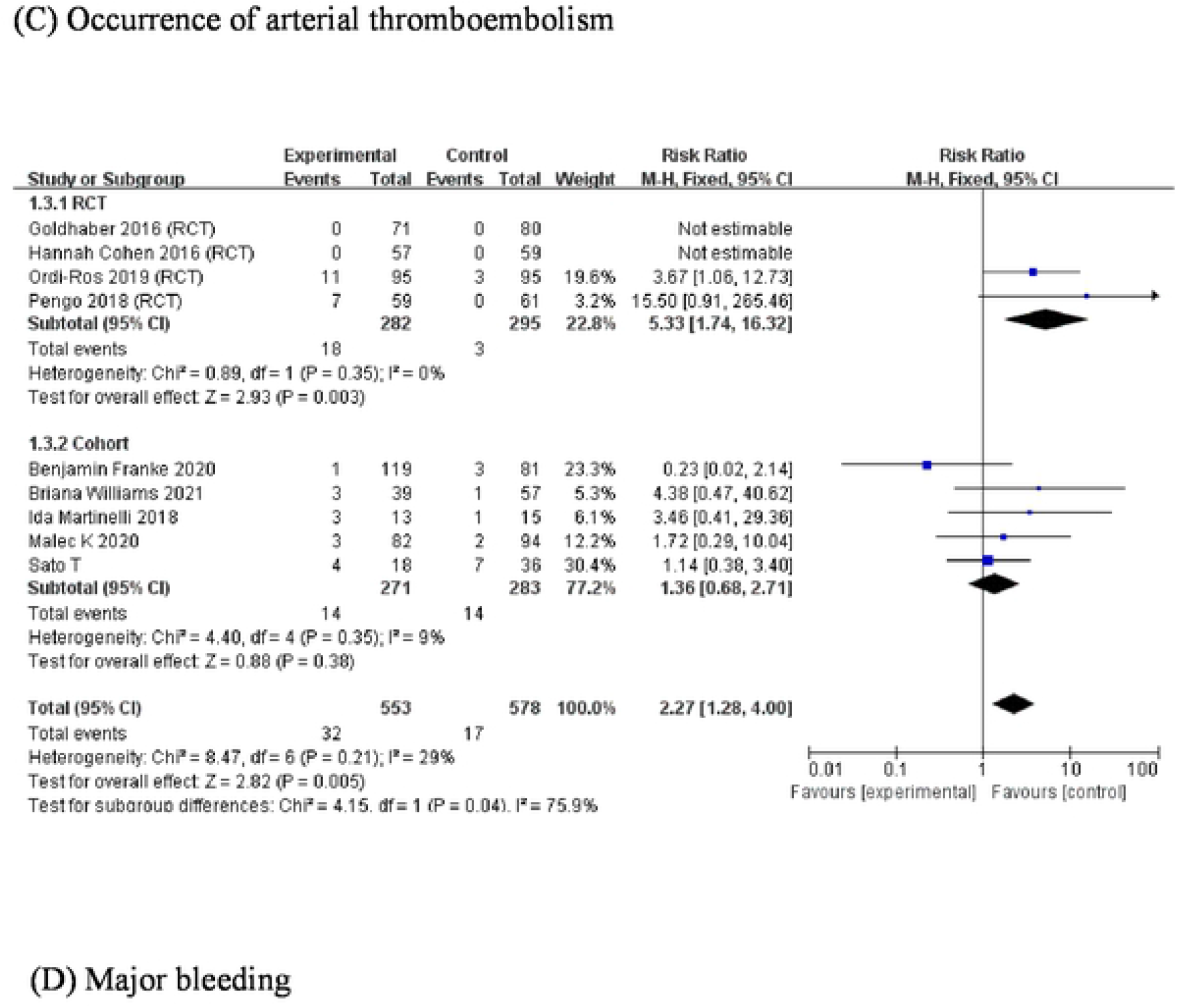
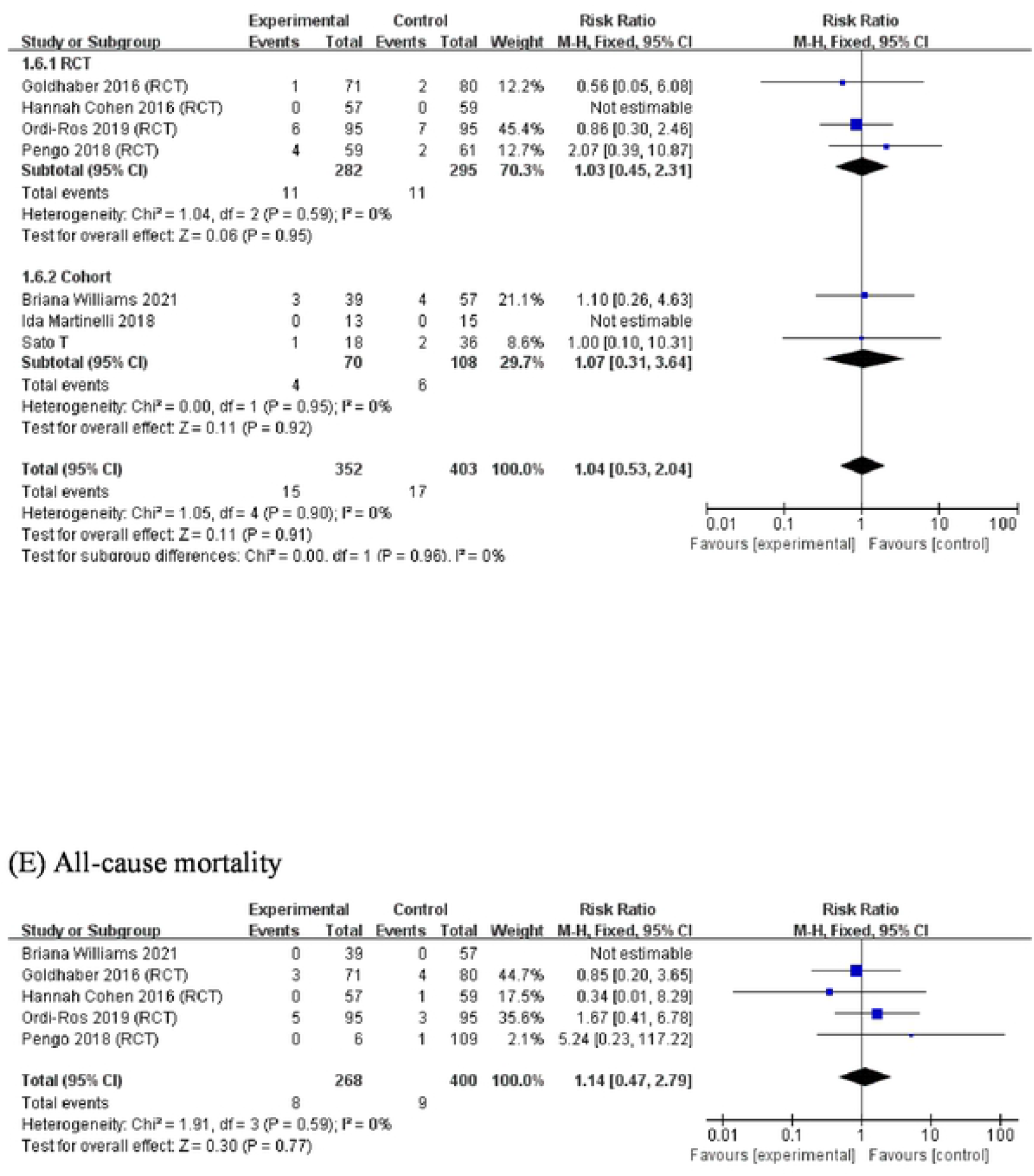
Forest plot for the efficacy and safety of DOACs in overall APS patients. (A) Occurrence of thromboembolic events. (B) Occurrence of venous thromboembolism. (C) Occurrence of arterial thromboembolism. (D) Major bleeding. (E) All-cause mortality.

The occurrence of major bleeding events and all-cause mortality was used to evaluate the safety of DOACs relative to VKAs in the treatment of APS. No significantly increased risk of major bleeding events (RR=1.04, 95% CI:0.53-2.04; I^2^=0%, P=0.91) was noted in the DOAC group compared to the VKA group. Both the RCT subgroup (RR=1.03, 95% CI:0.45-2.31; I^2^=0%, P=0.95) and the cohort subgroup (RR=1.07, 95% CI:0.31-3.64; I^2^=0%, P=0.92) demonstrated similar outcomes (Fig 3D). Pooled analyses (RR=1.14, 95% CI: 0.47-2.79; I^2^=0, P=0.77) found no amplified risk of all-cause mortality by DOACs in APS patients compared to VKAs (Fig 3E).

#### Comparing the efficacy and safety of DOACs versus VKAs for thromboprophylaxis in APS patients with different risk stratifications (Fig 4)

We explored the effectiveness and safety of DOACs versus VKAs in preventing thrombosis in APS patients with specified risk stratifications. The use of DOACs resulted in an increased risk of recurrent thrombosis (RR=3.65, 95% CI:1.49-8.93; I^2^=29%, P=0.005) in high-risk APS patients compared to VKAs; however, no substantial risk of recurrent thrombosis was found in the use of DOACs versus VKAs in the low-risk group (RR=1.65, 95% CI:0.72-3.81; I^2^=20%, P=0.24) (Fig 4A).

**Fig. 4.**
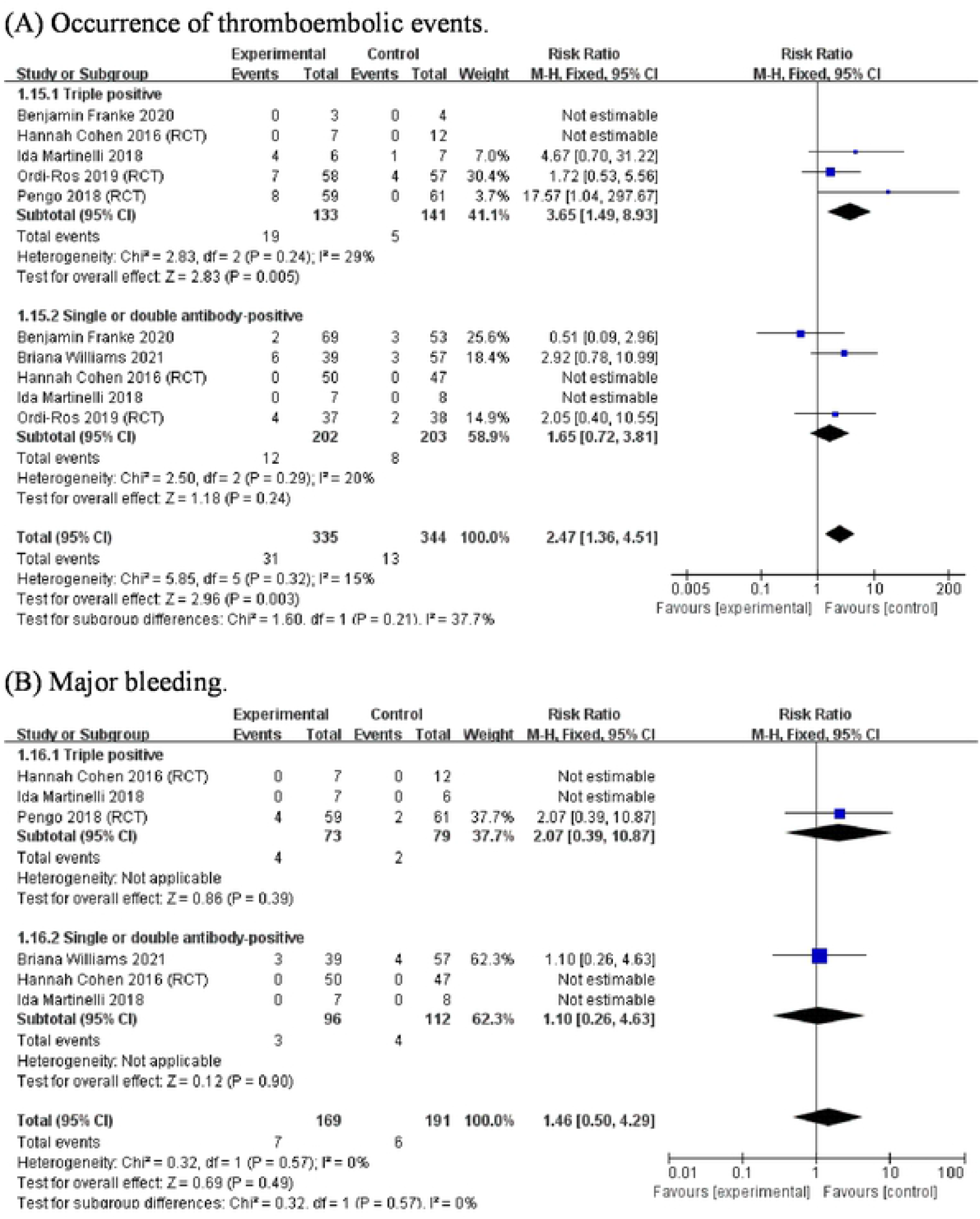
Subgroup analyses of the RRs of DOACs in treating triple-positive APS patients and non-triple-positive APS patients. (A) Occurrence of thromboembolic events. (B) Major bleeding.

There was no significantly augmented risk of major bleeding events in the high-risk subgroup when using DOACs compared to the application of VKAs (RR=2.07, 95% CI:0.39-10.87; P=0.39). This outcome was true for the low-risk subgroup too (RR=1.10, 95% CI:0.26-4.63; P=0.90) (Fig 4B).

The efficacy and safety of different DOACs could not be explored in APS patients with various stratifications due to the limited number of articles.

#### Comparing different DOACs to VKAs for thromboprophylaxis in overall APS patients (Fig 5)

In order to ascertain the efficacy and safety of various DOACs in the treatment of APS, we further examined the RRs for rivaroxaban, apixaban, and dabigatran-relayed recurrent thrombosis. Interestingly, only rivaroxaban correlated with a high risk of recurrent thromboses (RR=2.63; 95% CI, 1.56-4.42; I^2^ =0, P=0.0003) (Fig 5A) and a high risk of recurrent arterial thromboses (RR=4.52; 95% CI, 1.99-10.29; I^2^ =0, P=0.18) (Fig 5B) in patients with APS. The use of rivaroxaban incurred no heightened risk of recurrent venous thromboembolism relative to VKA utilization (RR=1.59, 95% CI: 0.79-3.18; I^2^=0, P=0.19) (Fig 5C). Patients treated with apixaban and dabigatran were not at higher risk of recurrent thrombosis, recurrent arterial thromboses and venous thromboembolism than those given VKAs.

**Fig. 5.**
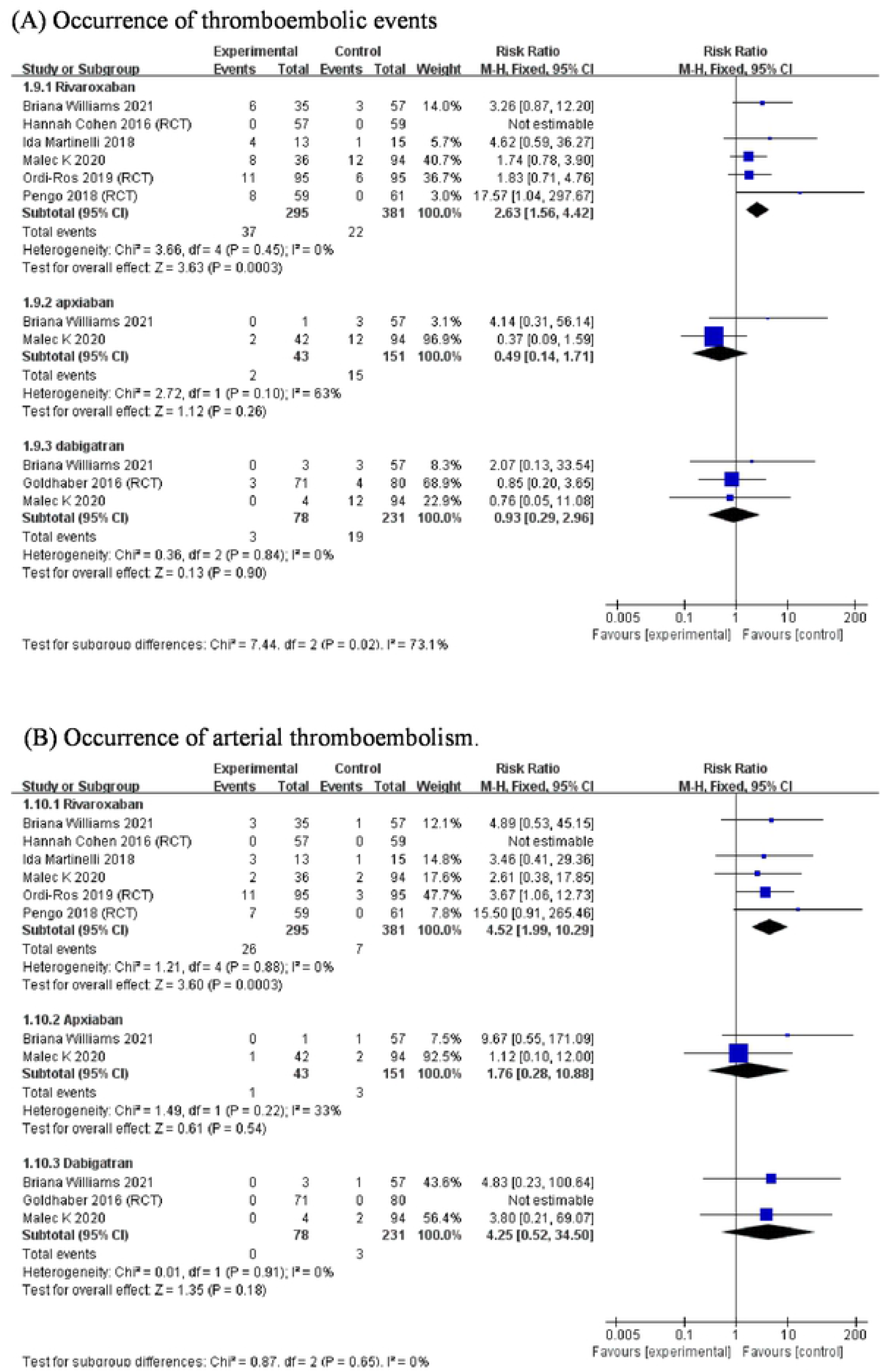
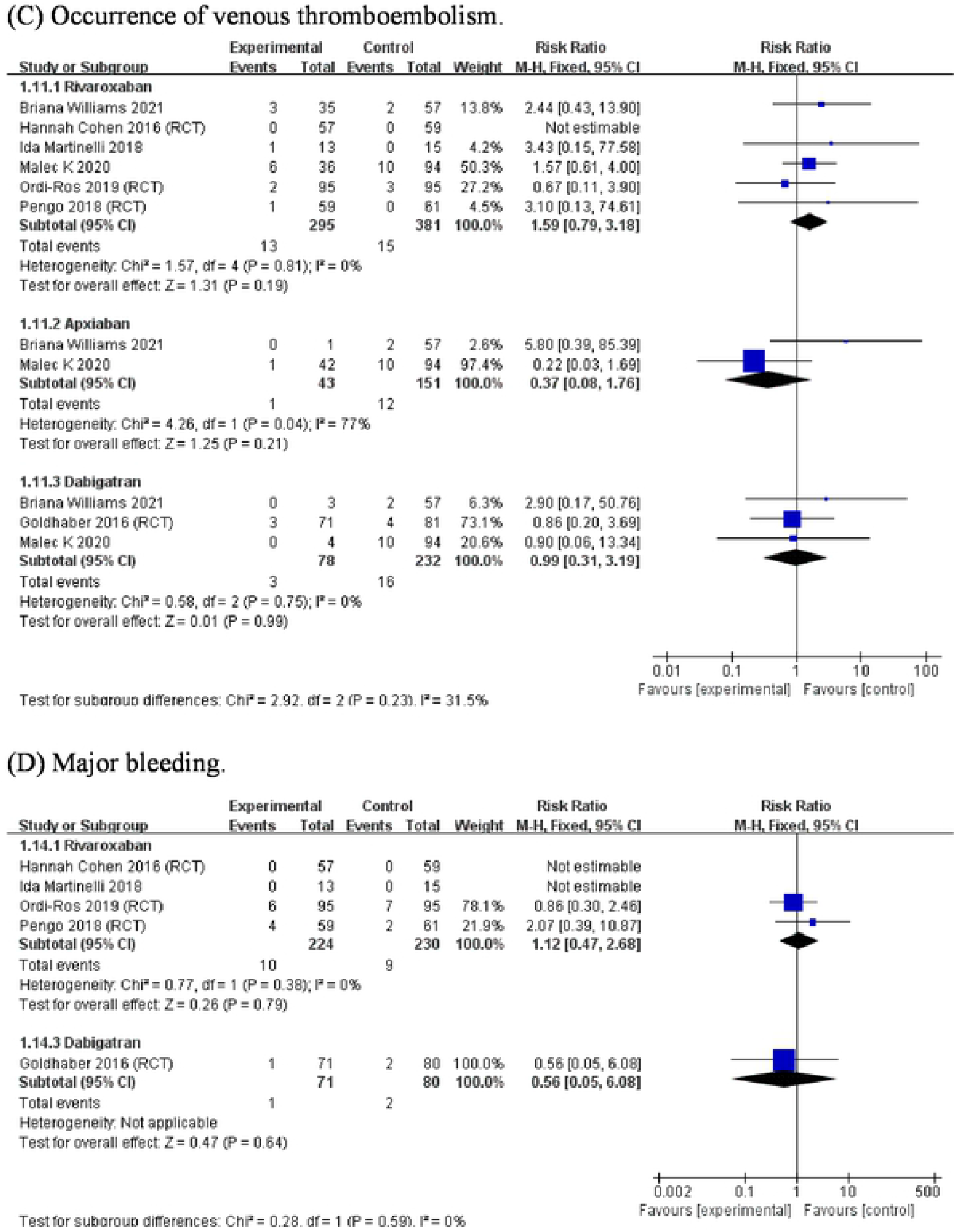
Subgroup analyses of the RRs for the efficacy and safety of different DOACs in treating patients with APS. (A) Occurrence of thromboembolic events. (B) Occurrence of arterial thromboembolism. (C) Occurrence of venous thromboembolism. (D) Major bleeding.

Additionally, rivaroxaban and dabigatran prompted no significantly amplified risk of major bleeding than the use of VKAs (Fig 5D).

#### Publication bias and heterogeneity analysis

Begg’s and Egger’s tests determined no significant publication bias (p=0.216) (Supplementary Figure 1). Sensitivity analyses to evaluate the robustness of the results by excluding one study after another and re-analyzing the data identified no influence by a particular publication. However, subgroup analyses indicated that study designs (RCT and cohort study), DOACs types, and risk stratification could cause heterogeneity.

## Discussion

Per our meta-analysis, the effectiveness of DOACs for thromboprophylaxis in high-risk APS patients is poorer the impact of VKAs; however, DOACs were comparable to VKAs in their efficiency in low-risk patients. Taking into account different APS patient types, rivaroxaban, which was the most representative drug in all included studies, statistically increased the incidence of arterial thromboembolism, but not venous thromboembolism, compared to VKAs; the other two DOACs (apixaban and dabigatran) displayed a similar tendency, but it was not statistically significant. In terms of safety, there was no statistical difference in the risk of significant bleeding and all-cause mortality in overall APS patients when using DOACs versus when taking VKAs.

Our meta-analysis innovatively explored the effectiveness and safety of DOACs versus VKAs in preventing thrombosis in APS patients with specified risk stratifications. We found that the incidence of thromboembolic events in patients with high-risk APS profiles in the DOAC group was nearly four-fold that in the VKA group, with statistical significance, which was inconsistent with the conclusion derived by a recent meta-analysis.[16] Dufrost et al.’s meta-analysis showed a trend towards a non-significant higher risk of recurrent thrombosis during treatment with DOACs compared to VKA use in APS patients with triple positivity. [16] Perhaps our addition of two recently published retrospective cohort studies[6,8] to our meta-analysis is responsible for this discrepancy. One of the two recent retrospective investigations found no thromboembolic events in the small minority of patients with triple positivity during follow-up. [6] The other inquiry, on the other hand, identified five of thirteen rivaroxaban users with recurrent thrombosis and one of fifteen patients in the VKA group with recurrent thrombosis; all the patients with recurrent thrombosis had triple positivity. [8] These findings provide a certain amount of evidence for guidelines recommending the use of warfarin but not DOACs to treat high-risk APS patients. [1,13,19] Given that our meta-analysis found no statistical significance in the difference in impact between DOACs and VKAs on the incidence of thromboembolic events in low-risk APS patients, DOACs could be an appealing therapeutic alternative thanks to its convenience and stability in this specified subgroup. More prospective studies or RCT trials must be scrutinized further to establish the viability of this conclusion.

DOACs differ in various ways, such as mechanisms and pharmacokinetics. [25] Rivaroxaban and apixaban are factor Xa inhibitors, while dabigatran is a thrombin (IIa) inhibitor. Rivaroxaban only needs to be taken orally once a day, whereas apixaban and dabigatran are taken twice a day. By comparing the efficacy and safety of different DOACs versus VKAs, this meta-analysis identified only rivaroxaban as displaying an unfavorable profile in the recurrence of thromboembolic events, especially arterial thrombosis, possibly because of the differences in anticoagulation strength required for arteriovenous thrombosis prevention. Investigations with animal models have shown that preventing arterial thrombosis requires a more potent inhibition of Xa and a higher dose of rivaroxaban than does venous thrombosis. [26] Because the use of DOACs does not need monitoring and their doses were not regularly adjusted, the therapeutic concentration in the rivaroxaban group may have been insufficient, particularly among patients with poor compliance who tended to skip medication periodically. However, subjects in the VKA group who failed to reach treatment goals (INR, 2 to 3) were excluded, with such a situation an uncommon occurrence. Besides, the presence of a different VKA mechanism that inhibits the synthesis of coagulation factors II, VII, IX, and X involved in vitamin K could also possibly explain the inferiority of rivaroxaban. The pooled results in our meta-analysis demonstrating apixaban and dabigatran’s non-inferior profiles compared to VKAs must be verified further with more RCTs because the current sample sizes from the limited articles are too small.

In several diseases that require anticoagulation, such as venous thromboembolic event (VTE) and nonvalvular atrial fibrillation, DOACs have displayed a favorable risk-benefit profile against hemorrhage events compared to VKAs. [27-29] In this meta-analysis, unexpectedly, the risk of major bleeding increased, but without statistical significance, in overall APS patients in the DOACs (particularly rivaroxaban) group compared to the VKA group, matching findings by the two recently published meta-analyses. [16,17] However, this phenomenon should be interpreted cautiously because APS patients with a warfarin INR goal range other than 2.0-3.0 were left entirely out of the RCTs and observative studies included in our meta-analysis. A previous meta-analysis proved that the risk of bleeding events in atrial fibrillation patients with excessive anticoagulation (INR > 3) was significantly higher relative to the situation in patients who maintained the recommended INR of 2 to 3. [30] Therefore, in a real-world experience, DOACs may still represent an attractive alternative treatment option for APS patients looking to mitigate the risk of bleeding, especially those with low-risk; however, this theory must be further examined.

Our study has several limitations. First, the pooled sample sizes are small. Second, three of the included four RCTs trials were open-label designed trials with risks of performance and selection biases because they had no blinded participants and personnel to intervene. This very bias possibly also exists in the six included cohort studies. Third, in order to expand the pooled sample sizes, we jointly analyzed RCTs and observational studies; however, these two investigation types differ in methodology, which may have affected the pooled results, as determined by the subgroup analyses based on the experimental design procedures of included articles (Fig 3C). Fourth, the number of studies included in the meta-analysis, especially in some subgroup analyses, was limited; therefore, publication biases may not have been detected because of the relatively lower power. Hence, our conclusions are not robust; further attempts to establish certainties on the touched-on issues are recommended.

In conclusion, different intensities of anticoagulation strategies must be specified for APS patients with different risk stratifications. This meta-analysis favors the use of VKAs to treat high-risk APS patients and DOACs for patients with lower-risk forms of APS. Furthermore, while rivaroxaban did not perform well versus VKAs, research on the application of DOACs for anticoagulation in APS patients must continue.

## Data Availability

All relevant data are within the manuscript and its Supporting Information files.

## Ethical approval

This systematic review and meta-analysis does not require ethical approval.

